# Socioeconomic status and immune aging in older US Adults in the Health and Retirement Study

**DOI:** 10.1101/2022.03.21.22272721

**Authors:** Eric T. Klopack, Bharat Thyagarajan, Jessica D. Faul, Helen C. S. Meier, Ramya Ramasubramanian, Jung Ki Kim, Eileen M. Crimmins

## Abstract

**Background:** Life course socioeconomic and demographic factors including educational attainment, race and ethnicity, and childhood (SES) are very powerful predictors of large inequalities in aging, morbidity, and mortality. Immune aging, including accumulation of late-differentiated, senescent-like lymphocytes and lower level of naïve lymphocytes, may play a role in the development of the age-related health inequalities. However, there has been little research investigating association between socioeconomic status and immune aging, particularly age-related changes in lymphocyte percentages.

**Methods:** This study used nationally representative data from more than 9000 US adults from the Health and Retirement Study to investigate associations between educational attainment, race and ethnicity, and childhood SES and lymphocyte percentages.

**Results:** Respondents with lower educational attainment, Hispanic adults, and those who had a parent with less than high school education had lymphocyte percentages consistent with more highly accelerated immune aging compared to those with greater educational attainment, non-Hispanic White adults, and respondents who had parents with a high school education, respectively. Associations between education, Hispanic ethnicity, and parents’ education and late differentiated senescent-like T lymphocytes (TemRA) and B cells were largely driven by cytomegalovirus (CMV) seropositivity.

**Conclusions:** Results suggest that CMV is a major driver of observed SES inequalities in immunosenescence and may therefore be an important target for interventions. Naïve T lymphocytes may be particularly affected by socioeconomic position and may therefore be of particular interest to research interested in inequalities in health and aging.

## Introduction

Age-related changes in immune function has been associated with increased risk cardiovascular disease, cancer, and other chronic conditions as well as reduced efficacy in fighting acute infections and reduced response to vaccines^1–5^. The aging immune system is characterized by lower percentages of naïve T and B lymphocytes, higher percentages of late differentiated lymphocytes, increases in cytotoxic (CD8^+^) relative to helper (CD4^+^) T cells, and decreases in naïve helper T cells relative to memory helper T cells. As the thymus involutes with age, fewer naïve cells are available to respond to novel viral and vaccine exposures^6^. Late differentiated cells accumulate over time and are less effective in viral response and other senescent cell and cancerous cell clearance and produce inflammatory cytokines and display a senescence-associated secretory phenotype^7–10^. Though other immune cells are involved in the immune aging, past research suggests that lymphocytes—and T lymphocytes in particular—are most affected by age-related immune changes^6,11^. Though age is a very powerful predictor of these changes^4^, there is substantial individual variation within age groups. Uncovering the sources of this variance is important for understanding and combating age-related health problems and functional decline.

A robust body of research links socioeconomic status (SES; e.g., wealth, education, income, occupational prestige) with various age-related health outcomes generally^12^ and with immune dysfunction particularly^13^. Indeed, past research suggests that lower income is associated with age-related cell type percentages^14^. Education may affect immune cell percentages by increasing exposure to health risks like social stressors that can accelerate immunosenescence^2,15^.

Lower educational attainment is also associated with increased risk of human cytomegalovirus infection (CMV) infection^13,16^. CMV is a common herpesvirus that is typically asymptomatic. CMV remains latent (i.e., once it is acquired, it is never cleared) and can be reactivated throughout life. This ongoing exposure to low levels of CMV infection results in an increasing proportion of the immune cells being dedicated to the management of CMV, leading to the exhaustion of naïve lymphocytes and the accumulation of memory T cells devoted to the virus (so called memory inflation)^8,17,18^. Thus, CMV infection is a strong determinant of cell percentages that characterize an aging immune system.

Race and ethnicity are highly related to SES in the US and are also robustly associated with age-related health problems^19^. Most past research on immune aging has utilized small or all White samples, making comparisons across race and ethnicity difficult. There has been mixed evidence from research comparing European and African populations^20^ and comparing White, Black, and Hispanic adults in the US^21^ regarding racial and ethnic differences in CD4^+^ and CD8^+^ T cell percentages, and to our knowledge, no research focusing on racial and ethnic differences in cell types specifically relevant to the immune aging (e.g., naïve and late differentiated T and B cells). However, there is good reason to expect that non-White and Hispanic US adults would have percentages of these cells indicative of accelerated immune aging. Black and Hispanic US adults have higher CMV seroprevalence and higher CMV antibody levels among CMV seropositive individuals^13,22^. Racial and ethnic minority groups in the US are structurally disadvantaged and therefore are exposed to more environmental toxins and social stressors and have limited access to socioeconomic opportunity^19^, all of which are associated with the immune age phenotype^23,24^. Racial and ethnic minorities are also exposed to particular stressors like racial discrimination that over time weather individuals^25^, leading to immune dysfunction and accelerated aging^26,27^. We therefore expect that non-White US adults will have lymphocyte percentages more typical of accelerated immune aging.

SES in childhood is also a major determinant of adult health, independent of adult SES^28–30^. Low childhood SES is associated with adverse health behaviors^31,32^, greater adversity and toxic stress^33^, lower adult SES attainment^34,35^, and may directly alter developmental trajectories toward a more accelerated aging phenotype^36,37^. Past research in small samples has found that early life adversity increases risk of CMV infection and T cell senescence^17^.

Though understanding the drivers of the immune age phenotype may be important for understanding health changes with age, relatively little research has investigated the roles of socioeconomic factors and race and ethnicity in immune aging. What research has been conducted on SES and immune age phenotype generally has utilized limited community samples. This study builds on this past research by utilizing nationally representative data on over 9000 US adults from the Health and Retirement Study. This study represents the first time that detailed immnophenotyping has been conducted in a large US population representative sample of older adults.

We investigated first whether educational attainment, race/ethnicity, and childhood SES (assessed by parental education) were associated with percentages of age-related lymphocytes: percentages of naïve CD4^+^ and CD8^+^ T cells, late differentiated (so called TemRA) CD4^+^ and CD8^+^ T cells, the CD4:CD8 cell ratio, the CD4 naïve:CD4 total memory cell ratio, naïve B cells, and late memory B cells. We expect higher educational attainment, being non-Hispanic White, and higher parental education to be associated with higher percentages of naïve lymphocytes, lower percentages of late differentiated (e.g., TemRA and late memory) lymphocytes, and higher CD4:CD8 and CD4 naïve:CD4 total memory cell ratios. We next examined whether these effects differed by CMV seropositivity. Finally, we examined potential interactions among these factors.

## Methods

### Sample

This study utilizes the Health and Retirement Study (HRS) 2016 Venous Blood Study (N = 9,934). HRS is a national panel survey of older Americans, which when weighted is designed to be representative of community dwelling older Americans older than 50. The VBS consisted of panel members interviewed before 2016 at least once who consented to a venous blood draw. 78.5% of eligible respondents consented and 82.9% of those respondents complete a blood draw. Sample characteristics are described in the results section. More information about this study is available elsewhere^38^. Absolute counts and percentages of 24 different types of immune cells were assessed using Multiparameter flow cytometry using the standardized protocol by the Human Immunology Project^39^ with minor modifications performed on an LSRII or a Fortessa X20 flow cytometer (BD Biosciences, San Diego, CA) More detailed methods for this sample are published elsewhere^38,40,41^.

### Measures

Lymphocyte percentages were assessed following the protocol described above. In the current study, we focus on the percentage of naïve CD4^+^ (CD4^+^/CD3^+^/CD19^-^/CD45RA^+^/CCR7^+^/CD28^+^) and CD8^+^ cells (CD8^+^/CD3^+^/CD19^-^/CD45RA^+^/CCR7^+^/CD28^+^), TemRA CD4^+^ (CD4^+^/CD3^+^/CD19^-^/CD45RA^+^/CCR7^-^/CD28^-^) and CD8^+^ cells (CD8^+^/CD3^+^/CD19^-^/CD45RA^+^/CCR7^-^/CD28^-^), the ratio of CD4^+^ to CD8^+^ T cells using the quotient of counts of these compartments, the ratio of naïve to total memory CD4^+^ T cells (sum of central memory (CD4^+^/CD3^+^/CD19^-^/CD8^-^/CD45RA^-^/CCR7^+^/CD28^+^), effector memory (CD4^+^/CD3^+^/CD19^-^/CD8^-^/CD45RA^-^/CCR7^-^/CD28^-^) and TemRA cells, naïve B cells (CD3^-^ /CD19^+^/IgD^+^/CD27^-^), and late memory B cells^42^ (CD3^-^/CD19^+^/IgD^-^/CD27^+^). Variables that showed substantial skewness (skewness > 2; viz., CD4^+^ TemRA, the CD4:CD8 ratio, CD4 naïve:CD4 total memory cell ratio, and late memory B cells) were natural log transformed to approximate a normal distribution. These variables were standardized to have a mean of 0 and variance of 1 to ease comparison.

CMV seropositivity was assessed using IgG antibodies to CMV using the Roche e411 immunoassay analyzer (Roche Diagnostics Corporation, Indianapolis, IN). Cutoffs were used to identify non-reactive (<0.5 COI; coded 0), borderline (0.5 to <1.0 COI; coded 0), and reactive (>1.0 COI; coded 1) individuals. Non-reactive and borderline groups were combined and were used as the reference group. More information is available elsewhere^38^.

Educational attainment was assessed by self-reported years of education. Respondents were categorized as 0-11 years (reference group), 12 years, 13-15 years, and 16 or more years.

Race/ethnicity was assessed by self-report. Respondents were categorized as non-Hispanic White (reference group), Hispanic, non-Hispanic Black, and non-Hispanic other race.

Parental low education was assessed using respondents’ reports of their parents’ educational attainment from the RAND HRS longitudinal file. If respondents reported on both parents, they were coded as 1 if either parent had less than high school education. If respondents only reported on one parent, they were coded as 1 if that parent had less than high school education. Respondents who only reported on their parents’ education at Wave 2H reported only whether their parent had attended school for 8 or more years. These respondents were assigned 7.5 years if less than 8 years and 8.5 is assigned if 8 or more. We also ran these models excluding participants who only reported on their parents’ education at Wave 2H. Results for these models were highly similar and had an identical pattern of significant results.

We control for age in years and sex (male as reference) in all models. The weighted sample had a mean age of 67.66 years, ranging from 50 to 107. The sample was 55.02% female.

### Plan of Analysis

Of the initial 9,934 respondents, 201 were excluded because they were cohort ineligible (e.g., were not a member of one of the birth cohorts represented in HRS) or were living in a nursing home. An additional 85 respondents were excluded because they were missing data on CMV seropositivity. 548 were excluded because they were missing on all flow cytometry measures in the current study. Sample weights, strata, and PSUs from the HRS tracker file were used to weight the sample. Respondents missing Venous Blood Study specific weights were assigned their core 2016 weight. The resulting sample size for this study is 9100. However, some individuals were missing data on specific cell types or predictor variables. To retain information, we used all respondents with complete data on each cell type for analyses using that cell type as the dependent variable. Additionally, there were too few participants in the non-Hispanic other race category to make meaningful comparisons, so those 277 participants were excluded in analyses with race and ethnicity as the focal predictors. E-table 1 shows final sample sizes for analyses for each model.

A series of generalized linear models were estimated regressing each cell type percentage/ratio on educational attainment or race/ethnicity, age and sex and then on educational attainment or race/ethnicity, CMV seropositivity, age, and sex. We then estimated a final set of models regressing each cell type percentage/ratio on race/ethnicity, education, CMV seropositivity, age, and sex. Because educational attainment and parental education highly covaried, we also estimated a model in which participants were categorized as high or low in each (not shown). This model produced a similar pattern of results. All analyses were conducted in R 4.1.1^43^ using the survey package^44^.

## Results

The weighted sample had a mean age of 68 years, ranging from 50 to 107. The sample was 55% female, 78.12 % non-Hispanic White, 9.74% non-Hispanic Black, 9.01% Hispanic, and 3.13% non-Hispanic other race. 14.23% had 0-11 years of education, 29.15% had 12 years of education, 25.32% had 13-15 years of education, and 31.30% had 16 or more years of education. 62.97% of participants were CMV seropositive.

Figure 1 shows the weighted percent of participants who were CMV positive by age and sociodemographic factors (also see e-table 2). Higher education was associated with lower CMV seropositivity in our sample. Participants with fewer than 12 years of education had the highest percentage of seropositivity before age 80 and the percentage of CMV seropositive respondents remained stable across age (Figure 1, Panel A). Higher educated participants tended to have lower seropositivity at all ages, and seropositivity increased with age. Non-Hispanic White respondents had lower CMV seropositivity at all ages compared to both non-Hispanic Black and Hispanic respondents (Figure 1, Panel B). CMV seropositivity was also associated with parents’ education such that participants whose parent(s) had less than a high school education had higher percentages of CMV seropositivity (Figure 1, Panel C). Finally, when considering education and race/ethnicity, CMV seropositivity tended to be lower for people with higher educational attainment across racial/ethnic groups (Figure 1, Panel D). To assess whether the association between educational attainment and CMV serostatus differed by race/ethnicity we estimated a model predicting CMV seropositivity with interactions between educational attainment and race/ethnicity (not shown), and no interaction term was significant.

**Figure 1.**
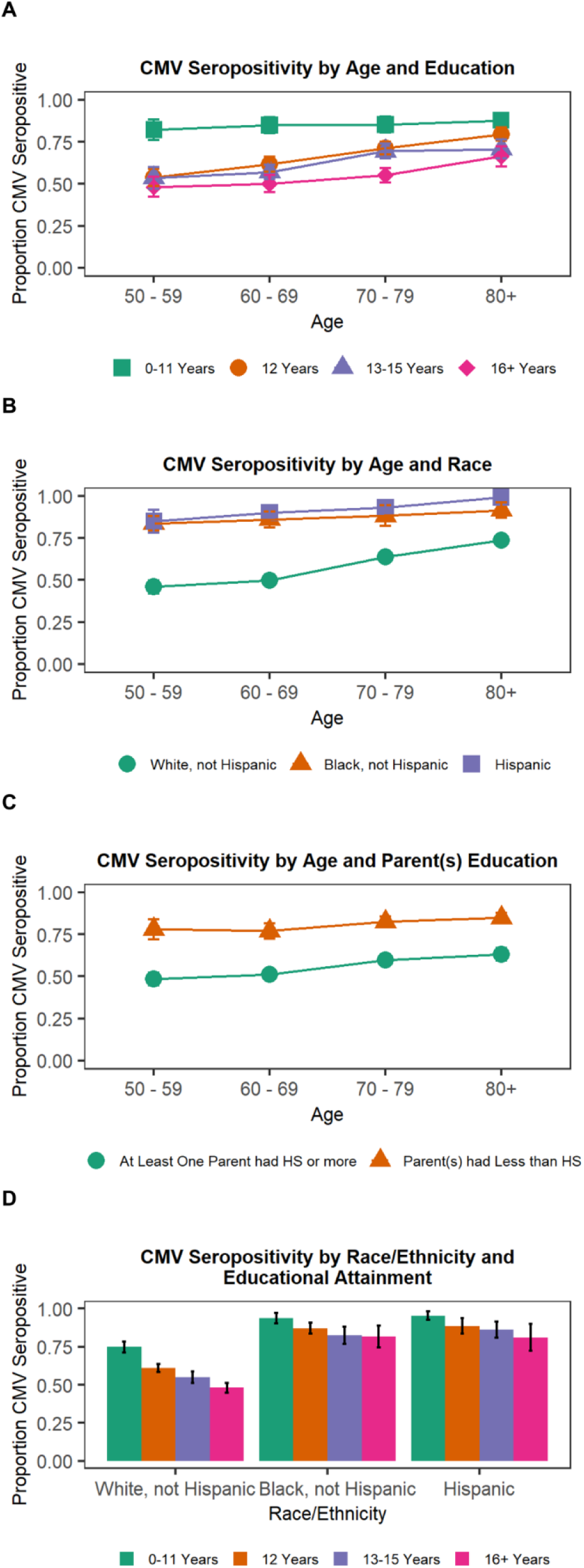
Percent CMV seropositive by SES variables and age Note: HS = high school; 95% confidence intervals shown.

Results indicating educational differences from models regressing cell type percentages and ratios on education and controls are shown in the left panel of Figure 2. Compared to participants with the lowest level of education, i.e. fewer than 12 years, having 12 years of education, 13-15 years of education, or 16 or more years of education was associated with a greater percentage of naïve CD4^+^ T cells, CD8^+^ T cells, higher CD4:CD8 T cell ratio and CD4 naïve:CD4 total memory cell ratios, and lower percentages of CD4^+^ and CD8^+^ TemRA and late memory B cells. Participants with 12 and 16 or more years of education had a greater percentage of naïve B cells compared to those with less than 12 years of education.

**Figure 2.**
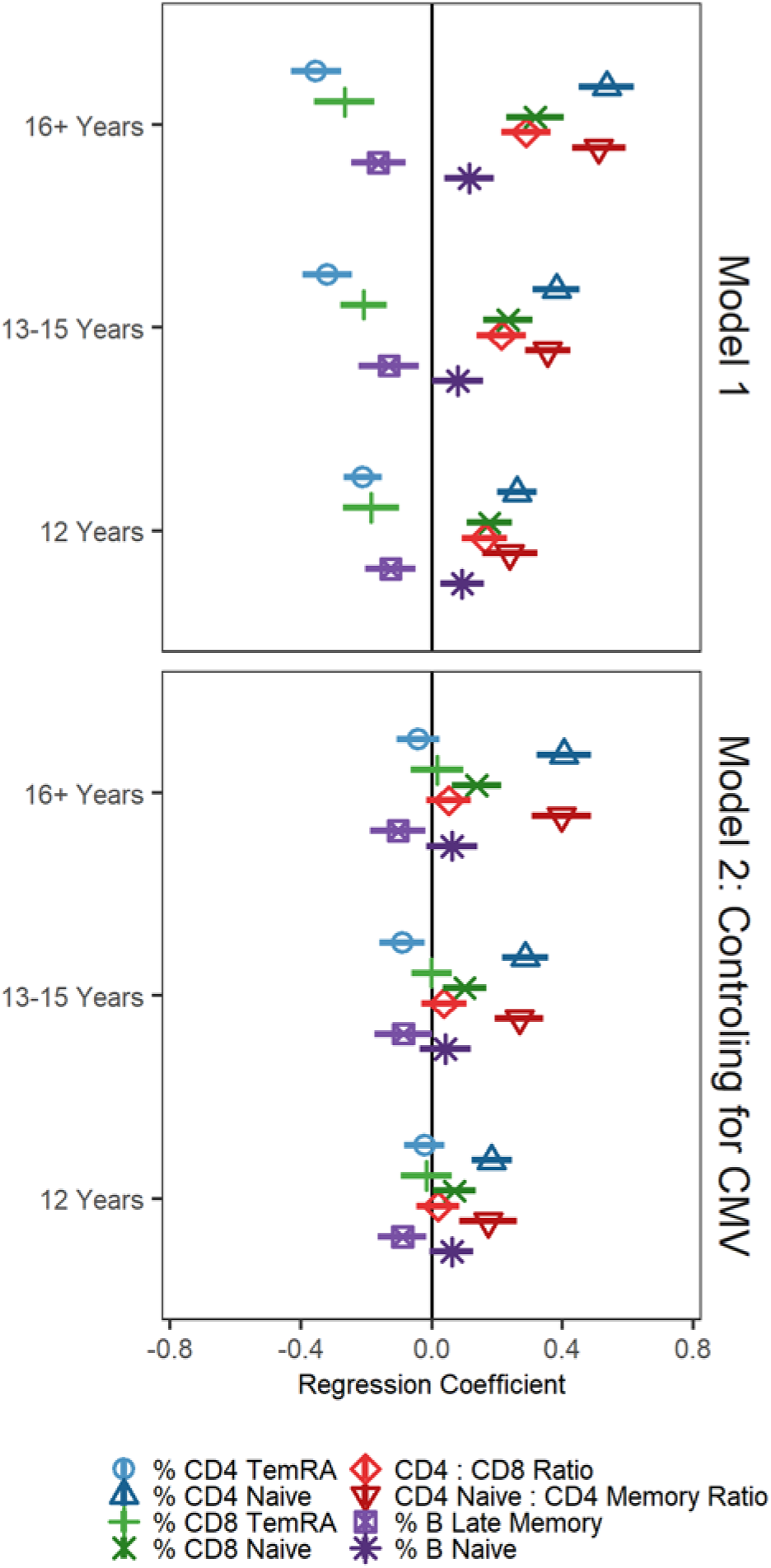
Educational attainment associated with cell type percentages and ratios Note: cell type percentages and ratios have been standardized to have a mean of 0 and standard deviation of 1; regression coefficients and 95% confidence intervals from GLMs regressing cell type percentages/ratios on educational attainment; reference group is less than 12 years of education.

The right panel of Figure 2 shows the same models additionally controlling for CMV seropositivity. Compared to having fewer than 12 years of education, having 12 years of education was still associated with a greater percentage of naïve CD4^+^, a higher CD4 naïve:CD4 total memory cell ratio, and fewer late memory B cells, but all other associations were attenuated and no longer statistically significant. Having 13-15 years of education was also associated with fewer CD8^+^ TemRA cells, a greater percentage of naïve CD4^+^ T and CD8^+^ T cells, and a higher CD4 naïve:CD4 total memory cell ratio but was no longer associated with CD4:CD8 ratio or with CD8^+^ TemRA and late memory B cells. Finally, having 16 or more years of education was associated with a greater percentage of naïve CD4^+^ T and CD8^+^ T cells, and a higher CD4 naïve:CD4 total memory cell ratio, and fewer late memory B cells.

Results from models regressing cell type percentages and ratios on race and ethnicity and controls are shown in the left panel of Figure 3. Compared to non-Hispanic White participants, non-Hispanic Black participants had fewer naïve CD4^+^ T cells, fewer naive B cells, and lower CD4:CD8 andCD4 naïve:CD4 total memory cell ratios. Black participants had a higher percentage of CD4^+^ TemRA cells, CD8^+^ naïve cells, CD8^+^ TemRA cells, and late memory B cells. Compared to non-Hispanic White participants, Hispanic participants had fewer naive CD4^+^ T and CD8^+^ T cells, lower CD4:CD8 and CD4 naïve:CD4 total memory cell ratios, and a higher percentage of CD4^+^ and CD8^+^ TemRA cells and late memory B cells.

**Figure 3.**
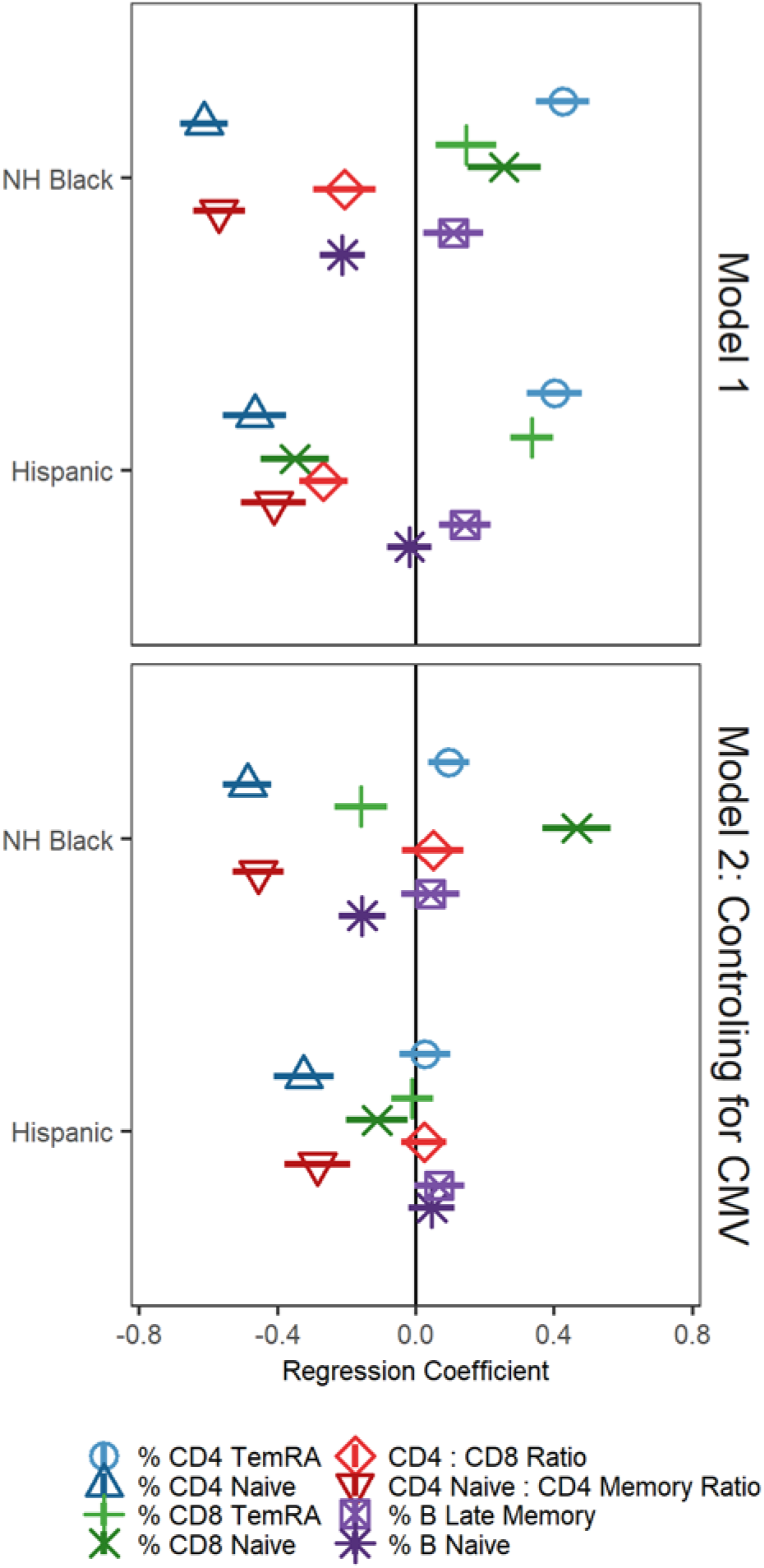
Race/ethnicity associated with cell type percentages and ratios Note: NH = non-Hispanic; cell type percentages and ratios have been standardized to have a mean of 0 and standard deviation of 1; regression coefficients and 95% confidence intervals from GLMs regressing cell type percentages/ratios on race/ethnicity; reference group is non-Hispanic White.

The right panel of Figure 3 shows the same models additionally controlling for CMV seropositivity. After controlling for CMV seropositivity, non-Hispanic Black participants no longer differed from non-Hispanic White participants on CD4:CD8 ratio or late memory B cell percentage. The non-Hispanic Black-White CD8^+^ TemRA cell percentage difference changed direction after controlling for CMV seropositivity. All differences between Hispanic participants and non-Hispanic White participants except on naïve T cell percentages and the CD4 naïve:CD4 total memory cell ratio were no longer statistically significant after controlling for CMV.

We further investigated the sign change in CD8^+^ TemRA cells for Black participants after controlling for CMV seropositivity noted in Figure 3 by looking at cell type percentages by race/ethnicity and CMV status (see e-table 3). Among CMV non-reactive and among CMV reactive individuals, non-Hispanic Black participants had a lower percentage of CD8^+^ TemRA cells compared to non-Hispanic White participants. These results are further discussed below.

Results of models regressing cell type percentages and ratios on parental education and controls are shown in the left panel of Figure 4. Participants who had at least one parent with less than high school education had a lower percentage of naïve CD4^+^ T, CD8^+^ T, lower CD4:CD8 T and CD4 naïve:CD4 total memory cell ratios, and higher percentages of CD4^+^ and CD8^+^ TemRA and late memory B cells. After controlling for CMV seropositivity, participants who had at least one parent with less than high school education had lower percentages of CD4^+^ and CD8^+^ naïve cells and lower CD4:CD8 T and CD4 naïve:CD4 total memory cell ratios compared to other participants.

**Figure 4.**
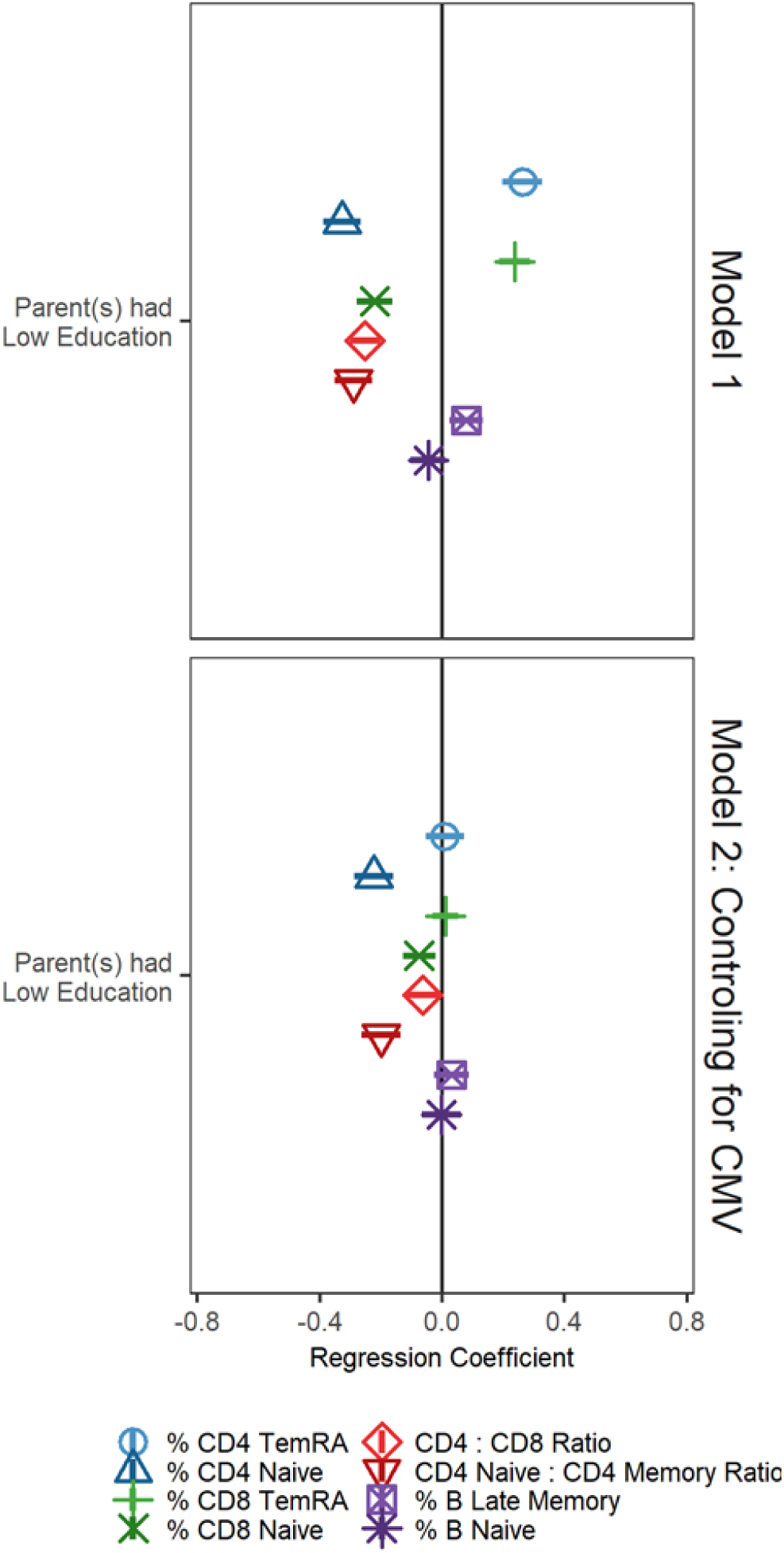
Parents’ Education associated with cell type percentages and ratios Note: cell type percentages and ratios have been standardized to have a mean of 0 and standard deviation of 1; regression coefficients and 95% confidence intervals from GLMs regressing cell type percentages/ratios on parents’ education; reference group is low parental education.

Next, all sociodemographic factors were included in one model, adjusting for covariates, to determine whether the estimates of associations were altered with mutual adjustment (shown in e-table 4). Controlling for childhood SES, race/ethnicity, and CMV serostatus, having 12 of years of education was associated with more CD4^+^ and CD8^+^ naïve cells compared to having less than 12 years of education. Having 12 years of education is no longer associated with the CD4 naïve:CD4 total memory cell ratio after including all covariates. Having 12 years of education is associated with a greater percentage of naïve B cells (a non-significant association in model shown in Figure 2) after including all covariates. Having 13-15 years of education was associated with Fewer CD4^+^ TemRA cells, more CD4^+^ and CD8^+^ naïve cells, and a higher CD4 naïve:CD4 total memory cell ratio. These associations were all also significant in results shown in Figure 2. Similar to results shown in Figure 2, having 16 or more years of education was associated with lower percentages of CD4^+^ and CD8^+^ naïve cells, and a higher CD4 naïve:CD4 total memory cell ratio, but was no longer with a lower percentage of late memory B cells. After controlling for all covariates, having 16 or more years of education was also associated with a higher percentage of naive B cells, a non-significant association in model shown in Figure 2.

Similar to results shown in Figure 3, compared to non-Hispanic White participants, non-Hispanic Black participants had more CD4^+^ TemRA cells, fewer CD4^+^ naïve cells, fewer CD8^+^ TemRA cells, more CD8^+^ naïve cells, a lower CD4 naïve:CD4 total memory cell ratio, and fewer naïve B cells. Similar to results shown in Figure 3, adjusting for childhood SES and educational attainment, compared to non-Hispanic White participants, Hispanic participants had fewer naïve CD4^+^ cells and a lower CD4 naïve:CD4 total memory cell ratio. Unlike results shown in Figure 3, Hispanic respondents no longer differed from White respondents in percentage of naïve CD8^+^ T cells. Also different from results shown in Figure 3, after controlling for all covariates, being Hispanic was associated with a higher CD4:CD8 ratio.

Similar to results shown in Figure 4, parental low education was associated with fewer CD4^+^ and CD8^+^ naïve cells, and a lower CD4:CD8 ratio adjusting for educational attainment and race/ethnicity. Unlike results shown in Figure 4, after controlling for all covariates, low parental education was not associated with the CD4 naïve:CD4 total memory cell ratio. Because educational attainment and parental education highly covaried, we also estimated a model in which participants were categorized as high or low in each (results not shown). Results from this model were similar to those in the model presented above, leading to the same conclusions.

Finally, we assessed interactions among these factors by regressing each cell type on all predictors and interactions among focal predictors (not shown). Non-Hispanic Black participants appear to benefit more from 13-15 and 16+ years of education in terms of naïve B cell percentage compared to non-Hispanic White participants. Non-Hispanic Black participants also appear to be less negatively affected by low parental education in terms of CD4^+^ naïve and B naïve cells compared to non-Hispanic White participants.

## Discussion

In this nationally representative sample of US adults over the age of 50, we found that cell type percentages and ratios indicative of immune aging were associated with SES and demographic factors. Higher educational attainment, non-minority status, and higher levels of parental education were associated less “aged” lymphocyte percentages. Though there has been little research on the association between immune aging and SES, these findings are consistent with evidence that income is associated with age-related immune change^14^ and that SES is associated with immune functioning broadly^13^. Our findings are also consistent with past research showing that early life SES is associated with CMV exposure and altered immune functioning^17^.

Controlling for CMV attenuated many of the associations with sociodemographic factors observed in this study, which underscores the importance of CMV in later life immune function. CMV infection is patterned by SES and race/ethnicity in the US^22^. Of note, individuals of low SES and minorities are more likely to acquire CMV earlier in life than high SES individuals and Non-Hispanic Whites^45^. Since the suppression of CMV requires substantial immunological resources, leading to memory inflation and the expansion of late-differentiated T cells^8,18^, having to control the virus for additional years may be relevant for the sociodemographic patterning of decline in immune function observed with aging^46^.

In this study, higher percentages of naïve T cells were consistently indicative of higher SES and non-minority status independent of CMV serostatus. It is possible that lifestyle factors associated with SES (e.g., diet and exercise) may slow thymic involution, leading to a relatively larger percentage of new naïve T cells being produced^7^. Thus, naïve T cells may be an immunological bellwether for socioeconomic and structural forces of interest to researchers interested in health inequalities, however further research is needed to refine this hypothesis.

Ratios among lymphocytes may be particularly useful for measuring age-related changes in immune cell percentages. Higher educational attainment was positively associated with CD4/CD8 ratio, indicating stronger immune function, however, all observed associations were attenuated and no longer statistically significant after adjusting for CMV. Thus, the differential prevalence of CD4^+^ and CD8^+^ T lymphocytes associated with socioeconomic position may be driven by differential exposure to and reactivation of CMV^22,47,48^. Alternatively, the ratio of CD4^+^ naïve to CD4^+^ memory cells was statistically significantly associated with educational attainment, race and ethnicity, and parental education after controlling for CMV seropositivity. Therefore, CD4^+^ naïve/CD4^+^ memory cell ratio may be particularly indicative of age-related immune phenotype that is affected by socioeconomic position.

Non-Hispanic Black-White differences in CD8^+^ naïve T cell percentages became more extreme after controlling for CMV and CD8^+^ TemRA cell differences changed direction. This was because non-Hispanic Black participants had a lower percentage of CD8^+^ TemRA cells and higher percentage of CD8+ naive T cells compared to non-Hispanic White participants within CMV seropositivity category. These differences were masked or reversed at the population level, suggesting this may be an example of Simpson’s paradox. That is, were it not for differential CMV exposure, Black US adults might have less accelerated immunosenescence compared to White US adults. Differential exposure to CMV may be a major driver of racial differences immunosenescence and potentially in racial health disparities, though further research is needed to confirm this. Interventions targeting exposure risk factors (e.g., number of individuals in the household, nativity outside of the US, region of residence, day care attendance, and sexual behaviors^22^) or potentially CMV vaccination may help in addressing racial health disparities. Future work on nutritional, psychological stress, inflammation, and morbidity and mortality prediction of immune aging is warranted.

This study has limitations. First, CMV seroprevalence and flow cytometry were only assessed at one time. Longitudinal assessments are needed to establish temporal ordering of CMV infection and age-related cell type percentages and ratios. Second, this study is limited to US adults 50 years and older. Further research in international samples with different educational systems and CMV seroprevalence profiles are needed to understand place-based SES and infection influences on later life immune function. Despite these limitations, this study has many strengths. The age-related cell type percentages and ratios examined expands on previous research into the immune risk profile^49,50^, immune parameters associated with mortality in very old individuals, to include age-related changes to immune function in a population representative study of older US adults. This study provides evidence that higher socioeconomic status may be protective against immunosenescence. Future research on nutritional, psychosocial stress, and inflammatory drivers of cell type percentages and ratios, as well as the relationship between cell type percentages and ratios and morbidity and mortality are warranted. Understanding the aging immune system may illuminate the biological processes underlying inequalities in age-related morbidity and mortality.

## Data Availability

All data are publicly available: https://hrs.isr.umich.edu/

