## Supporting Information for "Socioeconomic status and immune aging in older US Adults in the Health and Retirement Study"

**Online Supplement**

e-table 1. Sample Sizes for Each Model

| Dependent Variable | N for Models with Educational Attainment as the Focal Predictor | N for Models with Race and Ethnicity as the Focal Predictors | N for Models with Parents’ Education as the Focal Predictor | N for Models with All Predictors |
| --- | --- | --- | --- | --- |
| % CD4 TemRA | 9033 | 8754 | 8527 | 8205 |
| % CD4 Naive | 9033 | 8754 | 8527 | 8205 |
| % CD8 TemRA | 9034 | 8755 | 8528 | 8206 |
| % CD8 Naive | 9034 | 8755 | 8528 | 8206 |
| CD4:CD8 Count Ratio | 9078 | 8799 | 8569 | 8247 |
| CD4 Naive : CD4 Memory Ratio | 6708 | 6488 | 6333 | 6078 |
| % B Naive | 9056 | 8778 | 8550 | 8229 |
| % B Late Memory | 9056 | 8778 | 8550 | 8229 |

e-table 2. Descriptive statistics of the 2016 Health and Retirement Study with Flow Cytometry Data (N = 9100)

Panel A. Standardized cell type percentages and ratios and CMV seropositivity by educational attainment

|  | 0-11 Years | 12 Years | 13-15 Years | 16+ Years |  |
| --- | --- | --- | --- | --- | --- |
| % CD4 TemRA | 0.193 | -0.036 | -0.169 | -0.213 | a,b,c,d,e |
| % CD4 Naive | -0.243 | 0.019 | 0.149 | 0.271 | a,b,c,d,e,f |
| % CD8 TemRA | 0.176 | -0.059 | -0.156 | -0.207 | a,b,c,d,e |
| % CD8 Naive | -0.251 | -0.019 | 0.134 | 0.177 | a,b,c,d,e |
| CD4:CD8 Ratio | -0.152 | 0.018 | 0.090 | 0.143 | a,b,c,d |
| CD4 Naive : CD4 Memory Ratio | -0.222 | 0.009 | 0.120 | 0.249 | a,b,c,d,e,f |
| % B Naive | -0.079 | 0.031 | 0.048 | 0.072 | a,b,c |
| % B Late Memory | 0.105 | -0.023 | -0.032 | -0.061 | a,b,c |
| CMV Reactive | 0.853 | 0.658 | 0.603 | 0.524 | a,b,c,d,e,f |

Note: Cell type percentages and ratios have been standardized to have a mean of 0 and standard deviation of 1. Significant differences between groups (p < .05) are represented by: a = 0-11 years vs. 16+ years, b = 0-11 years vs. 13-15 years, c = 0-11 years vs. 12 years, d = 12 years vs. 16+ years, e= 12 years vs. 13-15 years, f = 13-15 years vs. 16+ years.

Panel B. Standardized cell type percentages and ratios and CMV seropositivity by race/ethnicity

|  | Non-Hispanic White | Non-Hispanic Black | Hispanic |  |
| --- | --- | --- | --- | --- |
| % CD4 TemRA | -0.169 | 0.242 | 0.210 | a,c |
| % CD4 Naive | 0.194 | -0.405 | -0.250 | a,b,c |
| % CD8 TemRA | -0.131 | -0.028 | 0.139 | a,b,c |
| % CD8 Naive | 0.040 | 0.351 | -0.222 | a,b,c |
| CD4 : CD8 Ratio | 0.092 | -0.099 | -0.150 | a,c |
| CD4 Naive : CD4 Memory Ratio | 0.177 | -0.386 | -0.224 | a,b,c |
| % B Naive | 0.052 | -0.144 | 0.059 | a,b |
| % B Late Memory | -0.041 | 0.063 | 0.095 | a,c |
| CMV: Reactive | 0.560 | 0.866 | 0.905 | a,c |

Note: cell type percentages and ratios have been standardized to have a mean of 0 and standard deviation of 1; significant differences between groups (p < .05) are represented by: a = non-Hispanic White compared to non-Hispanic Black, b = non-Hispanic Black compared to Hispanic, c = non-Hispanic White compared to Hispanic.

Panel C. Standardized cell type percentages and ratios and CMV seropositivity by parental education

|  | High Parental Education | Low Parental Education |  |
| --- | --- | --- | --- |
| % CD4 TemRA | -0.196 | 0.125 | a |
| % CD4 Naive | 0.193 | -0.120 | a |
| % CD8 TemRA | -0.229 | 0.173 | a |
| % CD8 Naive | 0.186 | -0.224 | a |
| CD4:CD8 Ratio | 0.137 | -0.130 | a |
| CD4 Naive : CD4 Memory Ratio | 0.162 | -0.092 | a |
| % B Naive | 0.065 | -0.042 | a |
| % B Late Memory | -0.043 | 0.040 | a |
| CMV: Reactive | 0.532 | 0.806 | a |

Note: cell type percentages and ratios have been standardized to have a mean of 0 and standard deviation of 1; significant differences between groups (p < .05) are represented by a.

e-table 3. Standardized cell type percentages and ratios by CMV status and race/ethnicity

|  | *CMV Non-Reactive* | | |  | | *CMV Reactive* | | | |  |
| --- | --- | --- | --- | --- | --- | --- | --- | --- | --- | --- |
|  | *Non-Hispanic White* | *Non-Hispanic Black* | *Hispanic* | |  | | *Non-Hispanic White* | *Non-Hispanic Black* | *Hispanic* |  |
| % CD4 TemRA | -0.760 | -0.566 | -0.490 | |  | | 0.302 | 0.363 | 0.280 | a |
| % CD4 Naive | 0.401 | -0.212 | 0.127 | |  | | 0.030 | -0.434 | -0.288 | a,b,c,d,e,f |
| % CD8 TemRA | -0.706 | -0.905 | -0.658 | |  | | 0.325 | 0.104 | 0.220 | a,d,e |
| % CD8 Naive | 0.444 | 0.977 | 0.572 | |  | | -0.281 | 0.257 | -0.301 | a,c,d,f |
| CD4:CD8 Ratio | 0.526 | 0.594 | 0.342 | |  | | -0.253 | -0.204 | -0.199 |  |
| CD4 Naive : CD4 Memory Ratio | 0.346 | -0.142 | 0.001 | |  | | 0.043 | -0.421 | -0.247 | a,b,d,e,f |
| % B Naive | 0.169 | -0.043 | 0.234 | |  | | -0.042 | -0.159 | 0.042 | a,c,d,e,f |
| % B Late Memory | -0.156 | -0.054 | -0.065 | |  | | 0.050 | 0.080 | 0.111 |  |

Note: Lymphocyte percentages and ratios are standardized to have a mean of 0 and variance of 1 to make comparisons easier, thus some values may be below 0 or above 1; significant differences between groups (p < .05) are represented by: a = non-reactive non-Hispanic White compared to non-reactive non-Hispanic Black, b = non-reactive non-Hispanic White compared to non-reactive Hispanic, c = non-reactive non-Hispanic White compared to non-reactive Hispanic, d = reactive non-Hispanic White compared to reactive non-Hispanic Black, e = reactive non-Hispanic White compared to reactive Hispanic, f = reactive non-Hispanic White compared to reactive Hispanic.

e-table 4. Estimates and p-values for associations between predictors and cell type ratios and percentages in the Health and Retirement Study

Panel A.

|  | % CD4 TemRA | | | | % CD4 Naïve | | | | % CD8 TemRA | | | | % CD8 Naïve | | | |
| --- | --- | --- | --- | --- | --- | --- | --- | --- | --- | --- | --- | --- | --- | --- | --- | --- |
|  | I | | II | | III | | IV | | V | | VI | | VII | | VIII | |
|  | b | p | b | p | b | p | b | p | b | p | b | p | b | p | b | p |
| Age | 0.011 | <0.001 | 0.005 | <0.001 | -0.002 | 0.196 | 0.000 | 0.955 | 0.031 | <0.001 | 0.025 | <0.001 | -0.037 | <0.001 | -0.033 | <0.001 |
| Gender (Female = 1) | 0.083 | <0.001 | -0.002 | 0.900 | 0.293 | <0.001 | 0.323 | <0.001 | -0.085 | 0.002 | -0.163 | <0.001 | 0.387 | <0.001 | 0.439 | <0.001 |
| Educational Attainment: 12 Years | -0.086 | 0.027 | -0.030 | 0.440 | 0.116 | 0.003 | 0.097 | 0.016 | -0.101 | 0.027 | -0.050 | 0.243 | 0.141 | <0.001 | 0.107 | 0.001 |
| Educational Attainment: 13-15 Years | -0.174 | <0.001 | -0.098 | 0.016 | 0.206 | <0.001 | 0.180 | <0.001 | -0.101 | 0.009 | -0.031 | 0.356 | 0.179 | <0.001 | 0.132 | <0.001 |
| Educational Attainment: 16+ Years | -0.179 | <0.001 | -0.050 | 0.227 | 0.306 | <0.001 | 0.261 | <0.001 | -0.131 | 0.009 | -0.013 | 0.754 | 0.248 | <0.001 | 0.170 | <0.001 |
| Race/Ethnicity: Black, not Hispanic | 0.374 | <0.001 | 0.103 | 0.003 | -0.532 | <0.001 | -0.439 | <0.001 | 0.079 | 0.108 | -0.168 | <0.001 | 0.344 | <0.001 | 0.509 | <0.001 |
| Race/Ethnicity: Hispanic | 0.271 | <0.001 | 0.012 | 0.790 | -0.306 | <0.001 | -0.216 | <0.001 | 0.214 | <0.001 | -0.022 | 0.556 | -0.198 | 0.001 | -0.041 | 0.426 |
| Parents' Low Education | 0.126 | 0.001 | -0.020 | 0.551 | -0.135 | <0.001 | -0.084 | 0.025 | 0.168 | <0.001 | 0.034 | 0.175 | -0.150 | <0.001 | -0.061 | 0.029 |
| CMV Seropositivity |  |  | 1.030 | <0.001 |  |  | -0.355 | <0.001 |  |  | 0.937 | <0.001 |  |  | -0.626 | <0.001 |
| Intercept | -0.859 | <0.001 | -0.992 | <0.001 | 0.005 | 0.967 | 0.051 | 0.667 | -2.123 | <0.001 | -2.244 | <0.001 | 2.190 | <0.001 | 2.270 | <0.001 |

Panel B.

|  | CD4:CD8 Ratio | | | | CD4 Naive : CD4 Memory Ratio | | | | % B Naive | | | | % B Late Memory | | | |
| --- | --- | --- | --- | --- | --- | --- | --- | --- | --- | --- | --- | --- | --- | --- | --- | --- |
|  | IX | | X | | XI | | XII | | XIII | | XIV | | XV | | XVI | |
|  | b | p | b | p | b | p | b | p | b | p | b | p | b | p | b | p |
| Age | -0.005 | 0.001 | 0.000 | 0.870 | 0.003 | 0.150 | 0.004 | 0.018 | -0.012 | <0.001 | -0.011 | <0.001 | 0.002 | 0.281 | 0.000 | 0.804 |
| Gender (Female = 1) | 0.211 | <0.001 | 0.275 | <0.001 | 0.259 | <0.001 | 0.293 | <0.001 | 0.089 | 0.002 | 0.103 | 0.001 | -0.034 | 0.195 | -0.050 | 0.057 |
| Educational Attainment: 12 Years | 0.099 | 0.010 | 0.056 | 0.136 | 0.094 | 0.087 | 0.081 | 0.143 | 0.089 | 0.028 | 0.080 | 0.046 | -0.097 | 0.038 | -0.087 | 0.059 |
| Educational Attainment: 13-15 Years | 0.129 | 0.004 | 0.070 | 0.077 | 0.175 | 0.001 | 0.151 | 0.003 | 0.078 | 0.089 | 0.066 | 0.151 | -0.102 | 0.063 | -0.088 | 0.103 |
| Educational Attainment: 16+ Years | 0.176 | <0.001 | 0.077 | 0.061 | 0.289 | <0.001 | 0.249 | <0.001 | 0.111 | 0.012 | 0.090 | 0.039 | -0.129 | 0.019 | -0.105 | 0.050 |
| Race/Ethnicity: Black, not Hispanic | -0.142 | 0.003 | 0.065 | 0.155 | -0.501 | <0.001 | -0.418 | <0.001 | -0.197 | <0.001 | -0.153 | <0.001 | 0.088 | 0.083 | 0.038 | 0.442 |
| Race/Ethnicity: Hispanic | -0.110 | 0.017 | 0.087 | 0.030 | -0.235 | <0.001 | -0.156 | 0.007 | 0.026 | 0.526 | 0.068 | 0.095 | 0.096 | 0.046 | 0.049 | 0.297 |
| Parents' Low Education | -0.182 | <0.001 | -0.071 | 0.031 | -0.118 | 0.004 | -0.076 | 0.052 | -0.009 | 0.805 | 0.015 | 0.671 | 0.024 | 0.468 | -0.004 | 0.915 |
| CMV Seropositivity |  |  | -0.781 | <0.001 |  |  | -0.312 | <0.001 |  |  | -0.167 | <0.001 |  |  | 0.189 | <0.001 |
| Intercept | 0.242 | 0.027 | 0.342 | 0.002 | -0.293 | 0.040 | -0.232 | 0.089 | 0.719 | <0.001 | 0.741 | <0.001 | -0.028 | 0.814 | -0.052 | 0.662 |
